# Inflammasome activation and pulmonary viral loads define two distinct clinical outcomes in COVID-19

**DOI:** 10.1101/2022.06.24.22276878

**Authors:** Keyla S.G. de Sá, Luana A. Amaral, Camila C.S. Caetano, Amanda Becerra, Sabrina S. Batah, Isadora M. de Oliveira, Letícia S. Lopes, Leticia Almeida, Samuel Oliveira, Danilo Tadao Wada, Marcel Koenigkam-Santos, Ronaldo B. Martins, Roberta R. C. Rosales, Eurico Arruda, Alexandre T Fabro, Dario S. Zamboni

## Abstract

COVID-19 has affected more than half a billion people worldwide, with more than 6.3 million deaths, but the pathophysiological mechanisms involved in lethal cases and the host determinants that determine the different clinical outcomes are still unclear. In this study, we assessed lung autopsies of 47 COVID-19 patients and examined the inflammatory profiles, viral loads, and inflammasome activation. Additionally, we correlated these factors with the patient’s clinical and histopathological conditions. Robust inflammasome activation, mediated by macrophages and endothelial cells, was detected in the lungs of lethal cases of SARS-CoV-2. An analysis of gene expression allowed for the classification of COVID-19 patients into two different clusters. Cluster 1 died with higher viral loads and exhibited a reduced inflammatory profile than Cluster 2. Illness time, mechanical ventilation time, pulmonary fibrosis, respiratory functions, histopathological status, thrombosis, viral loads and inflammasome activation significantly differed between the two clusters. Our data demonstrated two distinct profiles in lethal cases of COVID-19, thus indicating that the balance of viral replication and inflammasome-mediated pulmonary inflammation led to different clinical outcomes. We provide important information to understand clinical variations in severe COVID-19, a process that is critical for decisions between immune-mediated or antiviral-mediated therapies for the treatment of critical cases of COVID-19.

## Introduction

After the COVID-19 pandemic, coronaviruses became frequent viral agents of acute respiratory distress syndrome (ARDS). Between 2019 and 2021, there were more than 255,000,000 confirmed cases and 5,127,000 deaths caused by the severe acute respiratory syndrome coronavirus 2 (SARS-CoV-2) worldwide (WHO, 2020). Severe ARDS is characterized by major pulmonary involvement, respiratory distress, systemic thrombosis, and death (Batah and Fabro, 2021; Bösmüller et al., 2021; Polak et al., 2020). The activation of the NLRP3 inflammasome has been described in response to SARS-CoV-2 infections (Cama et al., 2021; Eisfeld et al., 2021; Junqueira et al., 2022; Lucas, 2020; Rodrigues et al., 2021; Sefik et al., 2022), and this process contributes to excessive inflammation and poor clinical outcome. The recruitment of immune cells to the lungs of infected individuals culminates in the excessive release of cytokines that cause structural damage to the lungs (Kommoss et al., 2020; Xu et al., 2020). Moreover, the major reported lung pathologies of severe COVID-19 include diffuse alveolar damage (DAD), acute fibrinoid organizing pneumonia (AFOP), and chronic interstitial pneumonia (Batah and Fabro, 2021; Bösmüller et al., 2021; Polak et al., 2020; Sauter et al., 2020). NLRP3 inflammasome activation also occurs in the lung parenchyma of these patients (Rodrigues et al., 2021; Toldo et al., 2021). However, it is still unknown how inflammasome activation promotes the exacerbated inflammatory process during SARS-CoV-2 infection and how it impacts viral replication and the development of important clinical characteristics associated with death in COVID-19 patients. In the present study, postmortem lung tissue samples from 47 fatal COVID-19 patients were examined for inflammasome activation and gene expression; in addition, these factors were correlated with the clinical conditions of the patients to understand the molecular mechanisms underlying the pathological processes that lead to the death of these patients. We demonstrated the presence of robust inflammasome activation in the lungs of patients infected with SARS-CoV-2 and defined the specific cell types that contribute most to inflammasome activation. An analysis of gene expression in pulmonary tissues allowed for the classification of COVID-19 patients into two different clusters: one cluster with patients who died with higher viral loads and reduced inflammatory profiles, which opposes the data from the other cluster. Our data establish that the magnitude of inflammasome activation contributes to the pathology of COVID-19 by balancing a thrombotic versus a fibrotic process that impacts the clinical outcome of the disease and patient death.

## Results

### Lethal cases of COVID-19 develop severe acute respiratory distress syndrome (ARDS) and higher inflammasome activation

We evaluated inflammasome activation in the lung parenchyma of 47 patients who died from SARS-CoV-2 infection from April to July 2020. These patients were infected with the ancestral strain of SARS-CoV-2 before the development of the COVID-19 variants of concern and before the use of vaccinations. As noninfected controls, we evaluated inflammasome activation in biopsies of the benign area of the lungs from patients who died due to lung adenocarcinoma (referred as uninfected controls). The data in **Table 1** show the demographic and clinical characteristics of the 47 patients. As reported in the early COVID-19 cases (that occurred in 2020), we found a high frequency of comorbidities, such as hypertension, obesity, diabetes, heart and lung disease. Furthermore, the patients exhibited altered values of CRP, D-dimers, LDH, creatinine, urea, AST, ALT, PT (INR), and blood glucose levels. The patients had an average illness time of 18 days, and the majority of the patients (76.59%) used MV, with an average of 12.38 days of use of mechanical ventilation. The patients had considerably altered PaO_2_ and PaO_2_/FiO_2_ characterizing a moderate to severe ARDS condition and consistent with the respiratory statuses. The histopathological analyses of COVID-19 patients demonstrated a high fibrotic phase of DAD, OP, and pneumonitis (**Table 1**). To analyze the histological features of patients’ lungs, we measured the lung parenchyma area in the histological sections to assess loss of airspace. In support of the data described above, we observed that COVID-19 patients had a higher parenchyma area (loss of airspace), suggesting that SARS-CoV-2 triggers robust inflammatory infiltration to the lung in severe cases of the disease (**Supplementary Fig. 1A-C**). These data are in agreement with histopathological findings showing an intense inflammatory process in patients who died from SARS-CoV-2 (Batah and Fabro, 2021; Hariri et al., 2021). To assess inflammasome activation in the lungs of COVID-19 patients, the lung parenchyma area and cell count were scanned in histological sections by using multiphoton microscopy. The parenchyma area was calculated by using ImageJ software, and inflammasome activation was microscopically scored by the presence of characteristic NLRP3 or ASC puncta/specks (Hauenstein et al., 2015). The parenchyma area was used to normalize all of the counts between the patients. Inflammasome activation was quantified by counting ASC and NLRP3 puncta in the lung tissue of these patients. We observed that patients with SARS-CoV-2 infection had robust inflammasome activation, as shown by the abundant presence of ASC and NLRP3 puncta in the lungs (**Supplementary Fig. 1D-G**). Representative images of ASC and NLRP3 puncta in the patients’ lungs are shown (**Supplementary Fig. 1E, G)**. Importantly, we observed ASC colocalization in nearly all of the scored NLRP3 puncta, thus confirming that these puncta structures that were abundantly found in the patient’s lungs are indeed the NLRP3/ASC inflammasome (**Supplementary Fig. 1H, I**). Collectively, these data indicate that COVID-19 patients have high inflammasome activation and that these patients evolved to a fibrotic phase of DAD, OP, and pneumonitis.

**Table 1.** COVID-19 patient characteristics

| **Demographic Characteristics** | Mean (±SD) or<br>N (%) |
| --- | --- |
| N | 47 |
| Sex |  |
| Female | 23 (48.93%) |
| Age (years) | 67.97 (15.05±) |
| Illness Time (days) | 18.08 (11.04±) |
| ICU | 37 (78.72%) |
| **Comorbidities** |  |
| Hypertension | 26 (55.32%) |
| BMI | 31.10 (±8.82) |
| Diabetes | 18 (38.29%) |
| History of smoking | 12 (25.53%) |
| Heart disease | 12 (25.53%) |
| Lung disease | 12 (25.53%) |
| Kidney disease | 7 (14.89%) |
| History of stroke | 7 (14.89%) |
| Autoimmune diseases | 2 (4.25%) |
| **Laboratorial findings** |  |
| CRP (mg/dL) | 11.40 (±8.49) |
| D-Dimers (μg/mL) | 4.91 (±4.46) |
| LDH (mmol/L) | 5.01 (±5.24) |
| Creatinine (mg/dL) | 2.22 (±1.4) |
| Urea (mg/dL) | 119.40 (±58.47) |
| AST (IU/L) | 104.42 (±90.74) |
| ALT (IU/L) | 72.12 (±55.14) |
| AST/ALT | 1.8 (±1.60) |
| PT (INR) | 1.81 (±2.13) |
| Albumin (g/dL) | 3.09 (±0.58) |
| Blood Glucose (mg/dL) | 194.72 (±94.60) |
| **Respiratory status** |  |
| Temperature ($^{\circ}$C) | 37.39 (±1.73) |
| Mechanical ventilation | 36 (76.59%) |
| Nasal-cannula oxygen | 11 (23.40%) |
| Intubated time (days) | 12.38 (±7.14) |
| P<sub>a</sub>O<sub>2</sub> (mmhg) | 74.01 (±24.63) |
| Venous saturation (S<sub>v</sub>O<sub>2</sub>) | 67.58 (±25.34) |
| P<sub>a</sub>O<sub>2</sub>/FiO<sub>2</sub><sup>#</sup> | 159.69 (±90.84) |
| A-a O<sub>2</sub> Gradient Value<sup>#</sup> | 283.22 (±208.31) |
| Respiratory Rate (mov/min) | 26.55 (±6.80) |
| **Histopathological findings** |  |
| Fibrosis (% of area) | 30.00 (±12.68) |
| Organizing Pneumonia (% of area) | 10.74 (±14.89) |
| Acute Fibrinous and Organizing Pneumonia (% of area) | 6.91 (±12.49) |
| Diffuse Alveolar Damage (% of area) | 17.87 (±21.05) |
| Pneumonitis (% of area) | 22.57 (±15.34) |
| Pulmonary infarction (% of area) | 3.404 (±9.618) |

\* Fisher's exact test
\*\* t-Test

### Macrophages and endothelial cells contribute to inflammasome activation in lethal cases of COVID-19

To comprehensively investigate the cell types operating during SARS-CoV-2 infection, we aimed to assess which cell types promote inflammasome activation in the lung of COVID-19 patients. We found abundant ASC and NLRP3 puncta in macrophages (CD64^+^), endothelial cells (CD34^+^), type 1 pneumocytes (PDPN^+^), and type 2 pneumocytes (SFTPC^+^) (**Figure 1A-H**). We scored ASC and NLRP3 puncta in CD64^+^, CD34^+^, PDPN^+^, and SFTPC^+^ and compared them with uninfected controls. We observed that in the lungs of COVID-19 patients, both ASC and NLRP3 puncta were significantly higher in macrophages (**Figure 1I, M**) and endothelial cells (**Figure 1J, N**) than in controls. In contrast, in the assessment of type 1 and type 2 pneumocytes, we found that ASC and NLRP3 puncta did not differ from uninfected controls, suggesting that these cells contribute less to overall inflammasome activation (**Figure 1K, L, O, P**). Analyzing the total numbers of macrophages, endothelial cells, type 1 pneumocytes, and type 2 pneumocytes in the lungs of patients who died from SARS-CoV-2 infection, we did not find differences in total counts of these cell types comparing COVID-19 and uninfected controls (**Supplementary Fig. 2A-D**). Together, these data demonstrate the main contribution of macrophages and endothelial cells but not pneumocytes, to inflammasome activation during SARS-CoV-2 infection.

**Figure 1.**
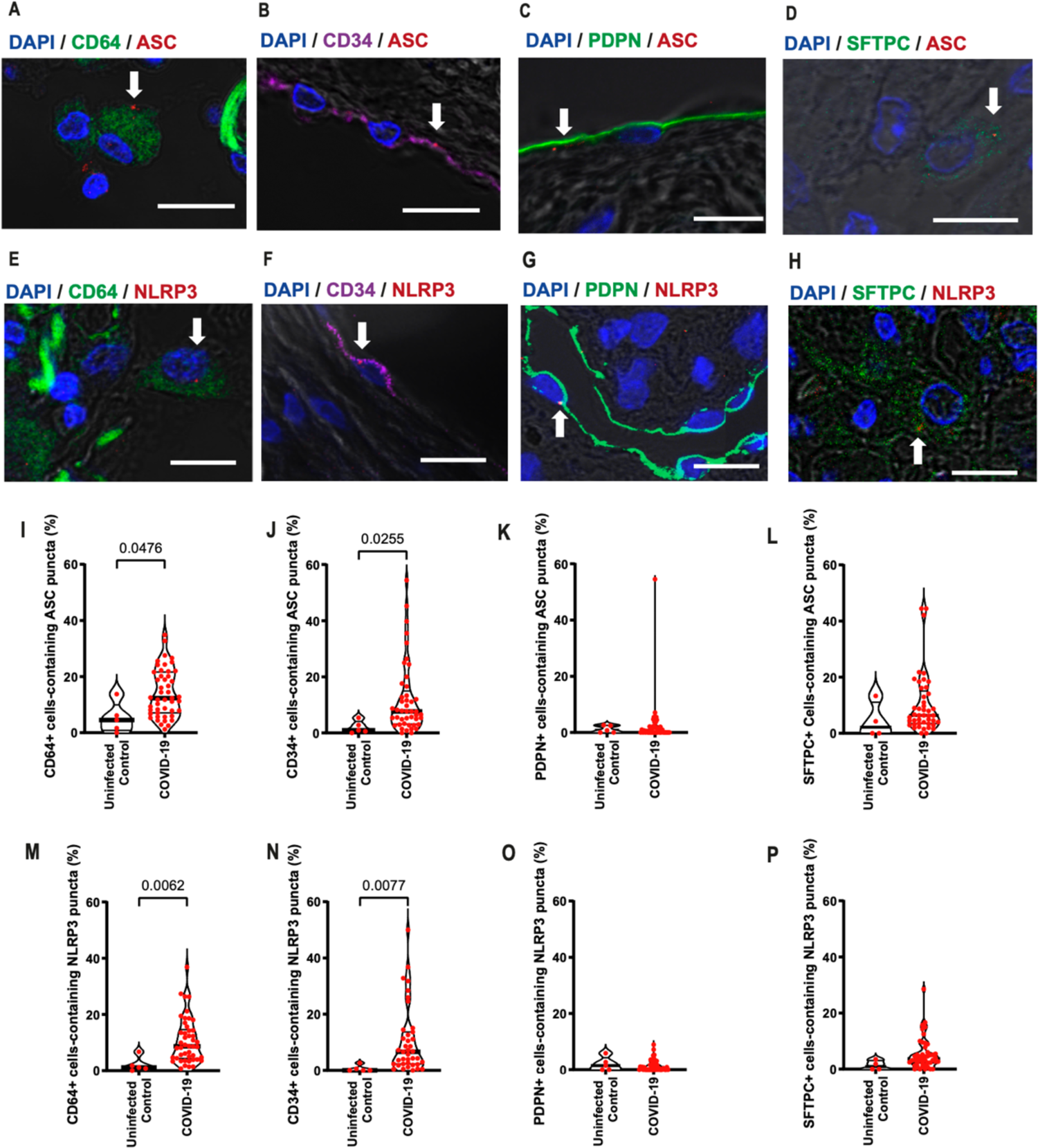
Macrophages and endothelial cells contribute to inflammasome activation in the lungs of COVID-19 patients. Multiphoton microscopy analysis of lung autopsies of 47 COVID-19 patients and 5 uninfected controls (benign area of the lungs from adenocarcinoma patients). Images of ASC (**A-D**) or NLRP3 (**E-H**) puncta (red, indicated by white arrows) in CD64^+^ (in green; **A, E**), CD34^+^ (in purple; **B, F**), PDPN^+^ (in green; **C, G**) and SFTPC^+^ (in green; **D, H**) cells from a lung autopsy of a COVID-19 patient. DAPI stains cell nuclei (blue). Scale bars 10 µm. The images were acquired by multiphoton microscopy using a 63x oil immersion objective and analyzed using ImageJ Software. (**I-P**) percentage of macrophages (CD64^+^), endothelial cells (CD34^+^), type I pneumocytes (PDPN^+^), and type II pneumocytes (SFTPC^+^) containing ASC (**I-L**) or NLRP3 (**M-P**) puncta. Each dot in the figures represents the value obtained from each individual. P-values are described in the figures comparing the indicated groups, as determined by Mann–Whitney test. Data are represented as violin plots with median and quartiles.

Subsequently, we performed immunohistochemistry analyses of the lung parenchyma to assess the expression of inflammasome components and observed variable expression of NLRP3, ASC, cleaved GSDMD, IL-1β, and caspase-1 in COVID-19 patients (**Supplementary Fig. 3A-E**). To assess which lung parenchyma cells expressed these inflammasome components, we used immunohistochemistry to evaluate the expression of ASC, NLRP3, caspase-1, cleaved GSDMD, and IL-1β on macrophages, endothelial cells, and type 2 pneumocytes. We also stained these tissues with anti-Spike of SARS-CoV-2 and found that virus-associated endothelial cells, macrophages, and type 2 pneumocytes expressed ASC, NLRP3, Caspase-1, and IL-1β (**Supplementary Fig. 3F-H**).

### Disease progression in fatal cases of COVID-19 occurs with decreasing viral load and increasing inflammasome activation

To assess whether inflammasome activation was associated with specific patient clinical conditions, we performed Pearson’s correlations in COVID-19 patients. Even though all of these patients died, we observed a strong negative correlation between viral load and the time of the disease (symptoms onset to death) (**Figure 2A, B**). In addition, we observed a positive correlation between the amount of NLRP3 and ASC puncta with the time of disease (**Figure 2C, D**), thus suggesting that, in general, whereas the viral load is reduced, inflammasome activation increases during hospitalization in these lethal cases of COVID-19. We also observed significant negative correlations between NLRP3 puncta and PaO_2_/FiO_2_ (**Figure 2E**), suggesting that inflammasome activation is related to an overall worsening pulmonary function. We detected no statistically significant correlations between NLRP3 or ASC puncta and viral load in the tissues (**Supplementary Fig. 4**).

**Figure 2.**
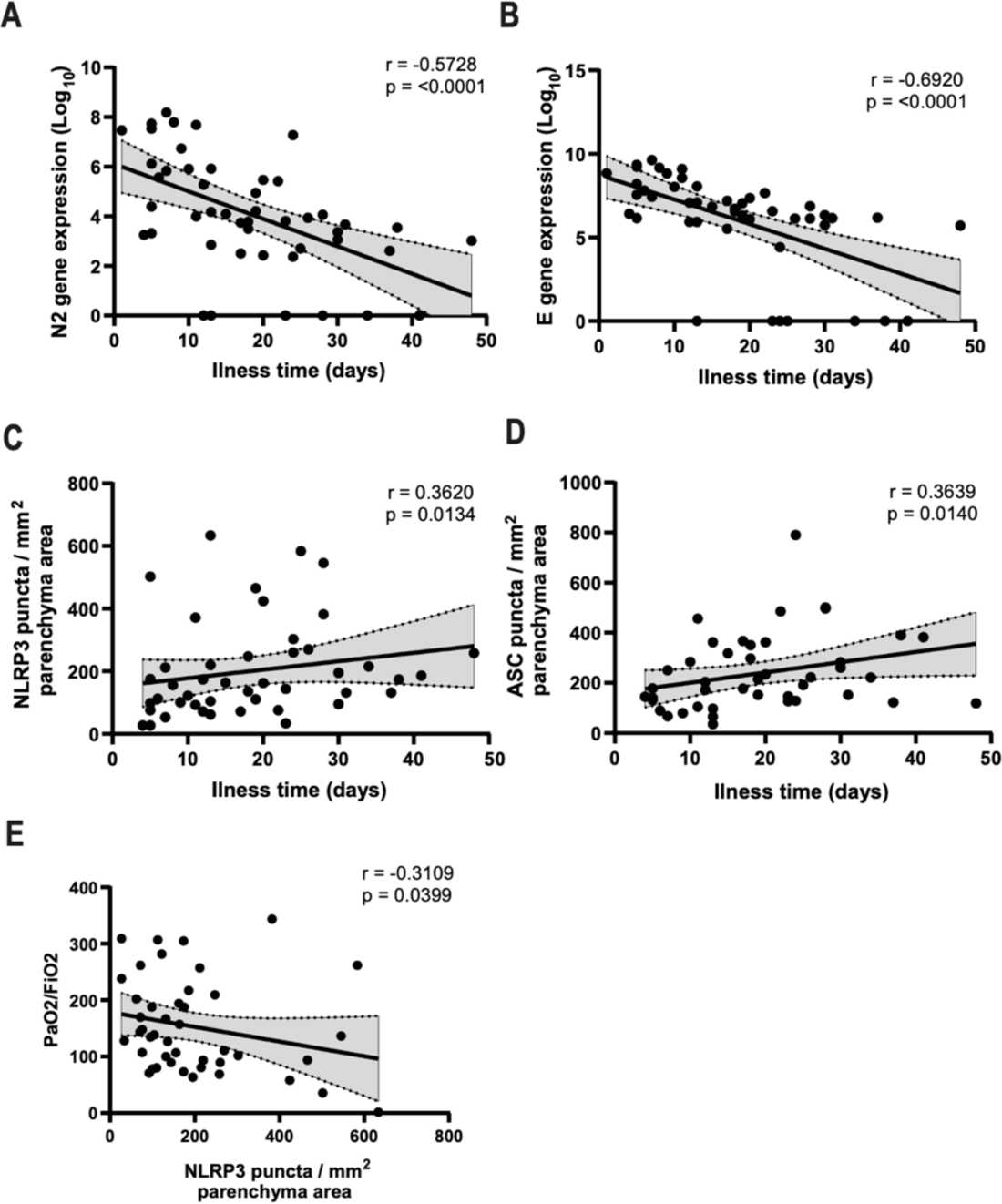
Inflammasome activation positively correlates with disease time, and viral load inversely correlates with disease time. Spearman correlation of pulmonary viral load, inflammasome activation and illness time in 47 fatal COVID-19 patients. (**A**) Correlation of viral N2 with illness time; (**B**) Correlation of viral E with illness time; (**C**) Correlation of NLRP3 puncta per parenchyma area with illness time; (**D**) Correlation of ASC puncta per parenchyma area with illness time; (**E**) Correlation of NLRP3 puncta per parenchyma area with PaO_2_/FiO_2_. r and P-values are indicated in the figures.

### Inflammasome activation and pulmonary viral loads define two distinct clinical outcomes in lethal cases of COVID-19

The imbalance of inflammatory and anti-inflammatory processes leads to an excessive release of cytokines into the systemic circulation with potentially deleterious consequences, including systemic inflammatory response syndrome (SIRS), circulatory shock, multiorgan dysfunction syndrome (MODS), and death (Sinha et al., 2020). Several studies have reported the association of SARS-CoV-2 with hyperinflammatory syndrome related to disease severity (Chen et al., 2021; Huang et al., 2005; Qin et al., 2020), and inflammasome activation may be associated with Cytokine Release Syndrome (CRS) (Cui and Zhang, 2020; Dolinay et al., 2012; Olajide et al., 2021). Thus, to correlate inflammasome activation with the hyperinflammatory profile observed in lethal cases of COVID-19, we analyzed the expression of genes related to the inflammatory process and inflammasome activation. We evaluated gene expression in the lungs of COVID-19 patients and observed that several inflammatory cytokine genes positively correlate with genes involved in inflammasome activation (**Supplementary Fig. 5A**). We did not observe a significant difference in the expression of these genes when we compared COVID-19 patients with uninfected controls (**Supplementary Fig. 5B-U**), possibly due to the large dispersion of data observed in COVID-19 patients.

Due to this dispersion and high variation in gene expression detected in COVID-19 patients, we performed an unsupervised heatmap constructed with data from relative gene expression and revealed the formation of two clusters in COVID-19 patients (**Figure 3**). Cluster 1 was characterized by a higher viral load and lower expression of inflammasome and inflammatory genes. In contrast to Cluster 1, Cluster 2 was comprised of patients who died with a lower viral load and increased expression of inflammasome and inflammatory genes (**Figure 3A**). To gain insights into key differences between these two clusters, we analyzed PaO_2_/FiO_2_ and A-a O_2_ gradient kinetics in these patients. We observed that patients in Cluster 2, who had overall increased inflammation, had worsening pulmonary function compared to those in Cluster 1 (**Figure 3B, C**). Basic demographic information stratified by cluster is provided in **Table 2**. Strikingly, patients who belonged to Cluster 2 had a long illness time (**Figure 3D**), worse pulmonary function (as indicated by the PaO_2_/FiO_2_ and A-a O_2_ gradients) (**Figure 3E, F**), greater inflammasome activation, as measured by NLRP3 puncta formation (**Figure 3G**), and increased area of pulmonary parenchyma, indicating loss or airways (**Figure 3H**). Images of the lung parenchyma of three representative patients from each cluster are shown (**Figure 3I-J)**. Importantly, we found that patients belonging to Cluster 2 had an overall lower viral load than patients from Cluster 1; this was quantified both by RT-PCR for N2 and E gene expression (**Figure 4A, B**), and by expression of Spike protein (**Figure 4C-E**). Next, we assessed radiological analyses of 15 patients from Cluster 1 and 31 patients from Cluster 2 by comparing the first and last chest X-ray images (CXR) and observed overall worsening pulmonary conditions in patients from Cluster 2 (**Supplementary Fig. 6A**). Representative images of CXR of two patients from Cluster 1, with Mild/Moderate pneumonia (stable conditions) (**Supplementary Fig. 6B**) and two patients from Cluster 2 with severe pneumonia (worsening) are shown (**Supplementary Fig. 6C)**. **Table 3** summarizes the general imaging evaluation findings for the patients in Cluster 1 and Cluster 2. Taken together, our data suggest the existence of two distinct groups of patients who succumbed to COVID-19, one group with lower viral loads, higher inflammation, and worse pulmonary conditions than the other group.

**Figure 3.**
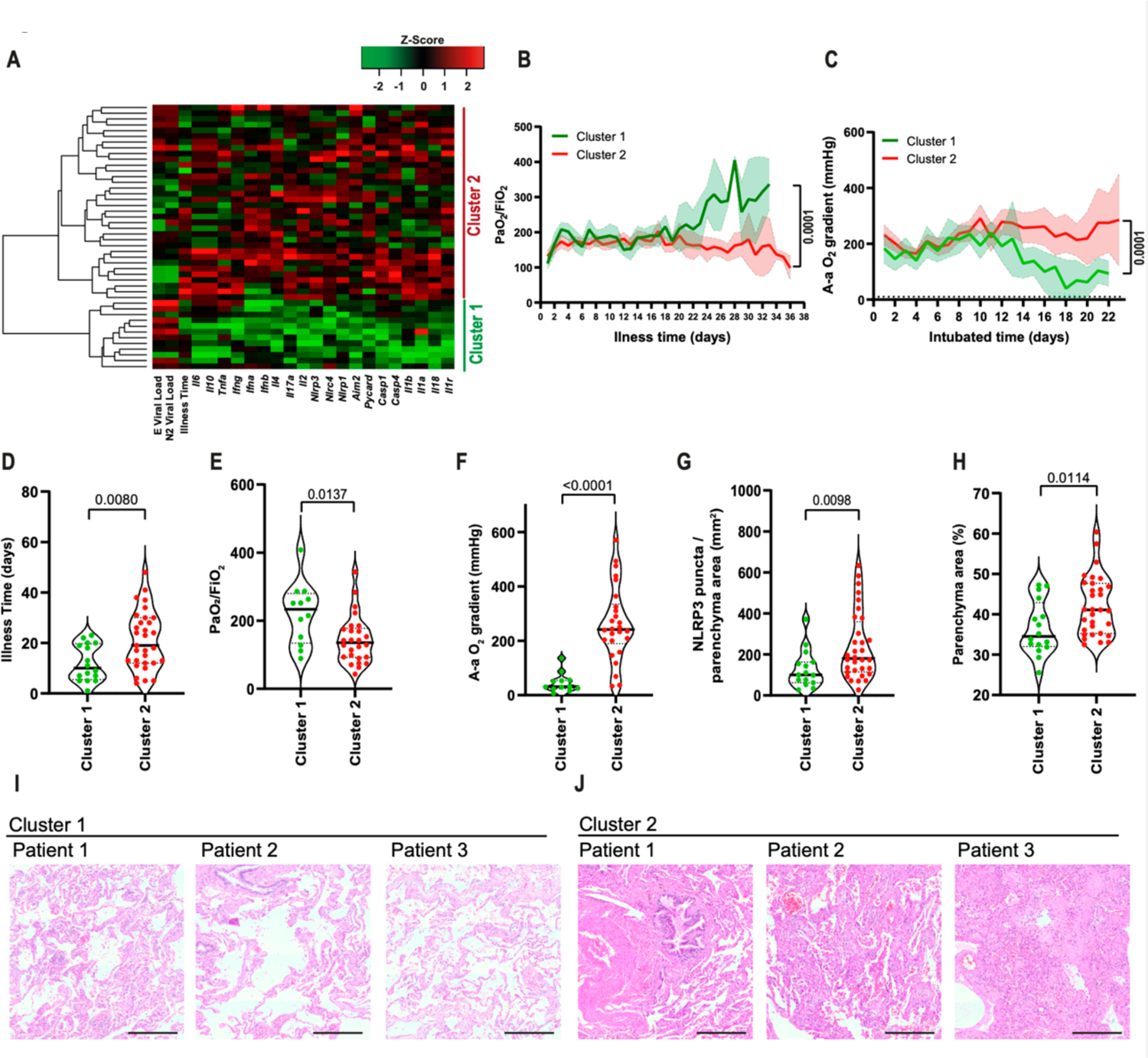
Pulmonary viral load and inflammatory gene expression define two patient clusters in lethal cases of COVID-19. (**A**) Heatmap of the mRNA expression of inflammasomes, inflammatory molecules/cytokines and viral N2 and E in lung autopsies of 47 COVID-19 patients. PaO_2_/FiO_2_ (**B**) and A-a O_2_ gradient (**C**) during disease development of patients from Cluster 1 (N=16, green) and Cluster 2 (N=31, red). *, *P* < 0.05 comparing the indicated groups, as determined by Area Under Curve test. (**D-H**) Analysis of Cluster 1 and Cluster 2 for illness time (**D**), PaO_2_/FiO_2_ (**E**), A-a O_2_ gradient (**F**), fibrosis (**G**), and NLRP3 puncta per lung parenchyma area (**H**). Each dot in the figure represents the value obtained from each individual. P-values are described in the figures comparing the indicated groups, as determined by Mann–Whitney test. Data are represented as violin plots with median and quartiles. (**I-J**) Representative H&E images of lung parenchyma of 3 patients of cluster 1 (**I**) and 3 patients of cluster 2 (**J**). Scale bars 200 µm.

**Figure 4.**
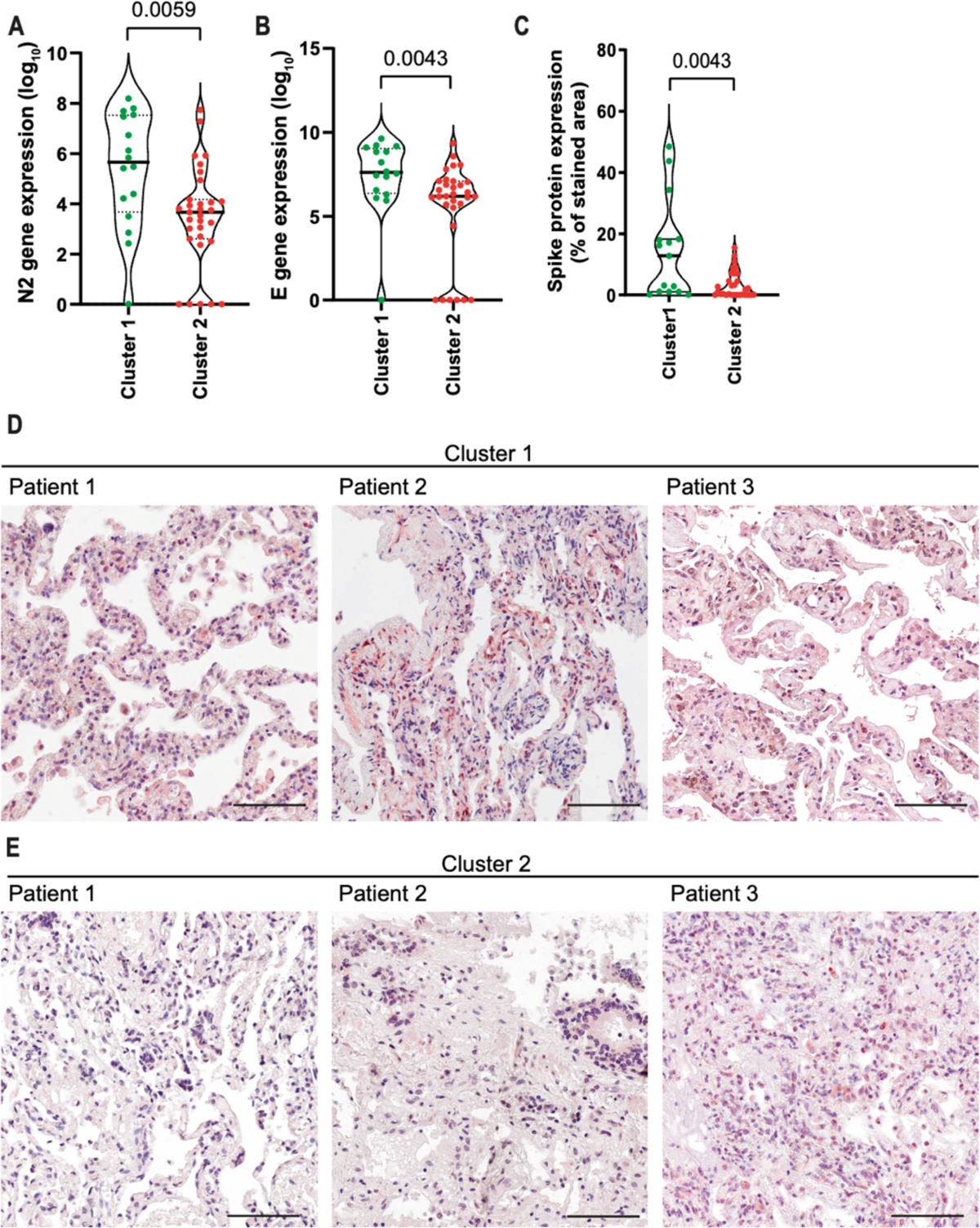
COVID-19 patients from Cluster 1 contain higher viral loads in the lungs than patients from Cluster 2. Quantification of viral N2 (**A**) and E (**B**) in lung autopsies of 47 COVID-19 patients from Cluster 1 (N=16) and Cluster 2 (N=31). (**C**) Quantification of the percentage of area stained for viral Spike protein in lung autopsies. Each dot in the figure represents the value obtained from each individual. P-values are described in the figures comparing the indicated groups, as determined by Mann–Whitney test. Data are represented as violin plots with median and quartiles. (**D-E**) Representative images of lung tissues stained for Spike (red) and Hematoxylin (blue). Scale bars 100 µm.

**Table 2.** COVID-19 patient characteristics

| Demographic Characteristics | Cluster 1<br>Mean (±SD) or<br>N (%) | Cluster 2<br>Mean (±SD) or<br>N (%) | P value |
| --- | --- | --- | --- |
| N | 16 | 31 | - |
| Sex |  |  |  |
| Female | 6 (37.50%) | 17 (54.83%) | 0.358* |
| Male | 10 (62.50%) | 14 (45.16%) |  |
| Age (years) | 70.81 (±10.34) | 66.5 (±17.46) | 0.369** |
| Illness Time (days) | 12.06 (±6.97) | 21.19 (±9.64) | 0.001** |
| ICU | 11 (68.75%) | 25 (80.64%) | 0.471* |
| **Comorbidities** |  |  |  |
| Hypertension | 10 (62.50%) | 16 (51.61%) | 0.546* |
| BMI | 28.39 (±6.92) | 33.21 (±8.52) | 0.057** |
| Diabetes | 7 (43.75%) | 11 (35.48%) | 0.999* |
| History of smoking | 4 (25.00%) | 8 (25.80%) | 0.999* |
| Heart disease | 4 (25.00%) | 8 (25.80%) | 0.999* |
| Lung disease | 3 (18.75%) | 9 (29.03%) | 0.505* |
| Kidney disease | 1 (6.25%) | 6 (19.35%) | 0.395* |
| History of stroke | 3 (18.75%) | 4 (12.90%) | 0.675* |
| Autoimmune diseases | 1 (6.25%) | 1 (3.22%) | 0.999* |
| **Laboratorial findings** |  |  |  |
| CRP (mg/dL) | 11.54 (±1.64) | 11.32 (±9.15) | 0.924** |
| D-Dimers (μg/mL) | 6.07 (±5.45) | 4.38 (±3.91) | 0.227** |
| LDH (mmol/L) | 6.17 (±6.42) | 4.41 (±4.52) | 0.280** |
| Creatinine (mg/dL) | 2.41 (±1.64) | 2.12 (±1.21) | 0.494** |
| Urea (mg/dL) | 108.36 (±53.92) | 125.09 (±60.75) | 0.358** |
| AST (IU/L) | 110.65 (±112.67) | 101.08 (±78.69) | 0.735** |
| ALT (IU/L) | 72.40 (±59.92) | 71.98 (±53.88) | 0.980** |
| AST/ALT | 2.1 (±2.47) | 1.6 (±0.92) | 0.319** |
| PT (INR) | 1.56 (±0.98) | 1.95 (±2.55) | 0.560** |
| Albumin (g/dL) | 3.24 (±0.65) | 3.01 (±0.54) | 0.251** |
| Blood Glucose (mg/dL) | 191.00 (±100.25) | 196.65 (±96.32) | 0.851** |
| Platelets ($10^3/\mu$L) | 173.70 (±97.83) | 273.10 (±128.20) | 0.010** |
| **Respiratory status** |  |  |  |
| Temperature ($^{\circ}$C) | 37.74 (±1.82) | 37.20 (±1.62) | 0.304** |
| Mechanical ventilation | 11 (68.75%) | 25 (80.64%) | 0.471* |
| Nasal-cannula oxygen | 5 (31.25%) | 6 (19.35%) | 0.471* |
| Intubated time (days) | 8 (±4.72) | 14.32 (±6.94) | 0.009** |
| P<sub>a</sub>O<sub>2</sub> (mmHg) | 75.54 (±8.06) | 73.27 (±1.69) | 0.135** |
| Venous saturation (S<sub>v</sub>O<sub>2</sub>) | 61.33 (±27.83) | 70.59 (±19.27) | 0.187** |
| P<sub>a</sub>O<sub>2</sub>/FiO<sub>2</sub><sup>#</sup> | 216.16 (±32.94) | 164.15 (±21.09) | <0.001** |
| A-a O<sub>2</sub> Gradient Value<sup>#</sup> | 40.20 (±101.48) | 237.58 (±68.05) | 0.006** |
| Respiratory Rate (mov/min) | 26.76 (±7.86) | 26.45 (±5.75) | 0.878** |
| PEEP | 9.63 (±5.08) | 9.62 (±4.86) | 0.9611** |
| **Histopathological findings, vascular status and Inflammasome activation** |  |  |  |
| Fibrosis (% of area) | 24.38 (±11.59) | 32.90 (±14.05) | 0.042** |
| Organizing Pneumonia (% of area) | 23.30 (±18.16) | 24.50 (±15.71) | 0.815** |
| Acute Fibrinous and Organizing Pneumonia (% of area) | 18.33 (±12.24) | 23.89 (±15.76) | 0.225** |
| Diffuse Alveolar Damage (% of area) | 34.00 (±24.03) | 33.33 (±20.23) | 0.920** |
| Pneumonitis (% of area) | 20.67 (±18.52) | 23.86 (±15.88) | 0.540** |
| Pulmonary infarction (% of area) | 26.67 (±14.14) | 20.00 (±14.14) | 0.132** |
| Disseminated Intravascular Coagulation | 8 (50.00%) | 5 (16.13%) | 0.0198* |
| NLRP3 puncta per parenchyma area (mm<sup>2</sup>) | 143.67 (±89.00) | 238.40 (±157.76) | 0.031** |
| Clearance Creatinine | 32.94 (±22.95) | 47.50 (±43.51) | 0.4380** |
| MDRD GFR | 41.58 (±30.03) | 44.15 (±36.90) | 0.9381** |
| Anticoagulation dose (mg) used close to death | 14.68 (±10.07) | 9.85 (±8.97) | 0.0125** |
| Glasgow scale | 7.18 (±5.70) | 6.61 (±5.23) | 0.7280** |
| SOFA score | 7.12 (±2.30) | 7.25 (±2.59) | 0.6828** |
| MELD score | 49.15 (±21.88) | 50.91 (±40.97) | 0.8113** |
| MODS | 10.57 (±3.45) | 10.39 (±3.16) | 0.9946** |

\* Fisher's exact test
\*\* t-Test

**Table 3.** Radiological Analyzes

|  | Cluster 1 | Cluster 2 | P value |
| --- | --- | --- | --- |
| **Initial CXR pattern** |  |  |  |
| No opacities | 1 (6.250%) | 3 (9.67%) | 0.9101 |
| Mild viral pneumonia | 2 (12.50%) | 3 (9.67%) |  |
| Moderate viral pneumonia | 8 (50.00%) | 12 (38.70%) |  |
| Severe viral pneumonia | 5 (31.25%) | 11 (35.48%) |  |
| Impaired analysis | 0 | 2 (6.45%) |  |
| **Last CXR pattern** |  |  |  |
| No opacities | 1 (6.25%) | 2 (6.45%) | 0.9971 |
| Mild viral pneumonia | 3 (18.75%) | 5 (16.12%) |  |
| Moderate viral pneumonia | 6 (37.50%) | 12 (38.70%) |  |
| Severe viral pneumonia | 4 (25.00%) | 8 (25.80%) |  |
| Impaired analysis | 2 (12.50%) | 4 (12.90%) |  |
| **Imaging evolution until death** |  |  |  |
| Improvement (fewer opacities) | 5 (33.33%) | 10 (32.25%) | 0.0364 |
| Stability | 7 (46.66%) | 4 (12.90%) |  |
| Worsening (more opacities) | 2 (13.33%) | 12 (38.70%) |  |
| Impaired analysis | 1 (6.66%) | 5 (16.12%) |  |
| **Disease Extent (Thoracic CT)** |  |  |  |
| 0 | 0 | 0 | 0.0363 |
| <25% | 4 (66.66%) | 1 (12.50%) |  |
| 25-50% | 0 | 3 (37.50%) |  |
| 50-75% | 1 (16.66%) | 3 (37.50%) |  |
| >75% | 1 (16.66%) | 1 (12.50%) |  |

### Differential regulation of fibrinolysis-related genes and growth factors dictate the induction of fibrosis or disseminated intravascular coagulation in fatal cases of COVID-19

Histopathological analyses of the lungs from lethal cases of COVID-19 belonging to Clusters 1 and 2 allowed us to detect an increased development of fibrosis in patients from Cluster 2 (**Figure 5A**, and **Table 2**). By contrast, patients belonging to Cluster 1 exhibited a markedly increased disseminated intravascular coagulation (**Figure 5B, and Table 2**), according to the parameters recommended by the International Society of Thrombosis and Hemostasis (ISTH) (Taylor et al., 2001). In support of our analyses demonstrating increased disseminated intravascular coagulation, we found reduced platelet counts in patients from Cluster 1 (**Figure 5C, and Table 2**), supporting that vascular dysfunctions impact clinical outcomes in patients belonging to Cluster 1.

**Figure 5.**
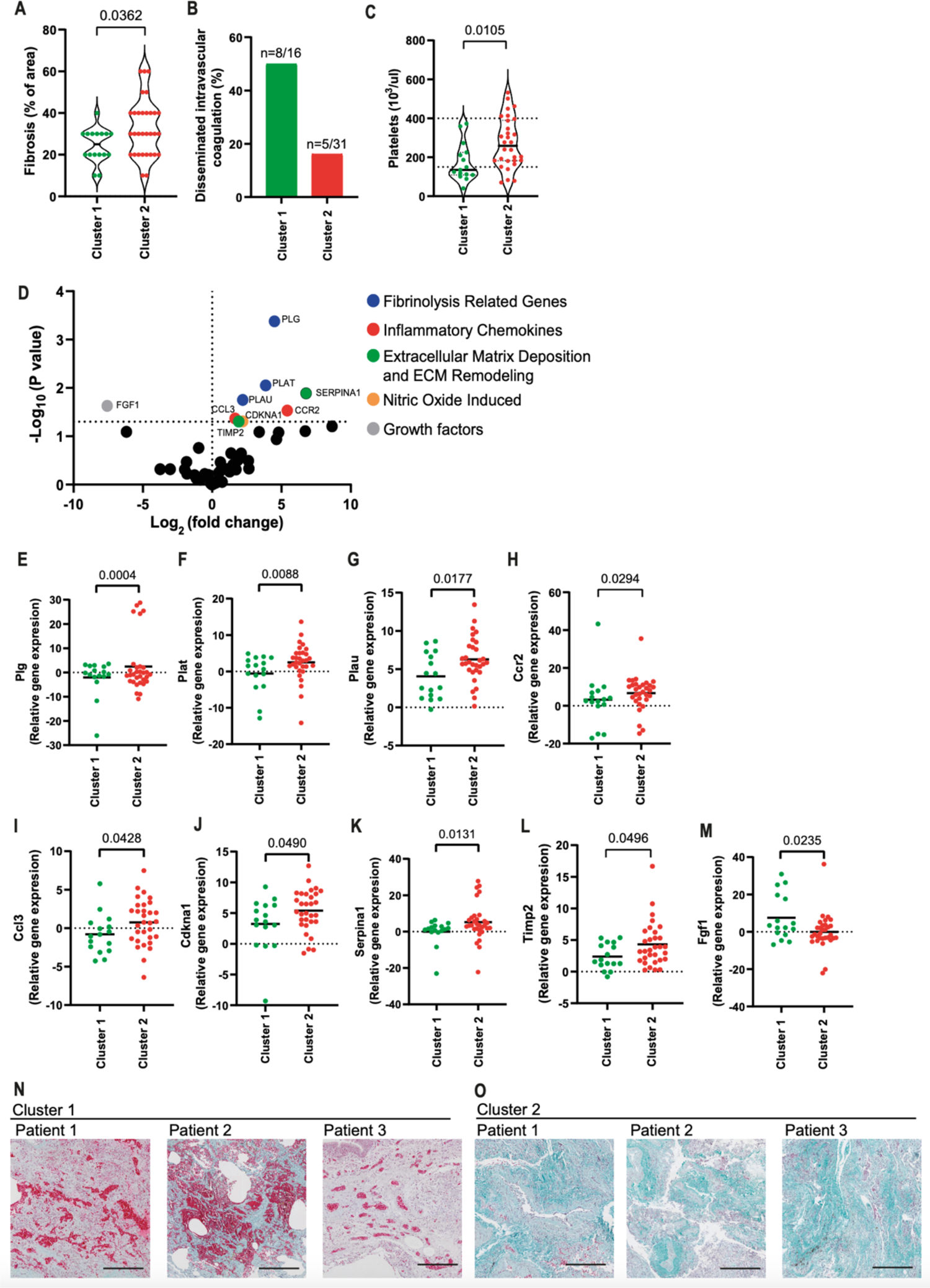
Fibrosis is increased in cluster 2 patients. (**A-B**) Histopathological analysis of pulmonary samples from Cluster 1 and Cluster 2 for the presence of fibrosis (**A**) and Disseminated Intravascular Coagulation (**B**). (**C**) Platelet counts by laboratory analyses of blood samples from COVID-19 patients on the last day of hospitalization. (**D**) Volcano plot analysis showing expression of fibrosis-related genes in lung autopsy of COVID-19 patients. Each dot in this figure represents a gene (average gene expression of 47 patient samples). Fold change Cluster 2/Cluster 1). The genes with statistically significant differences are indicated in color dots: *Plg* (**E**), *Plat* (**F**), *Plau* (**G**), *Ccr2* (**H**), *Ccl3* (**I**), *Cdkna1* (**J**)*, Serpina1* (**K**)*, Timp2* (**L**), and *Fgf1* (**M**). Each dot in the figures (**A, C, E-M**) represents the value obtained from each individual. P-values are described in the figures comparing the indicated groups, as determined by Mann–Whitney test. Data are represented as violin plots with median and quartiles. (**N-O**) Representative images of Masson-Goldner staining of lung parenchyma from three Cluster 1 patients (**N**) and three Cluster 2 patients (**O**). Shown are Nuclei (dark brown to black), Collagen (green/blue), Erythrocytes (bright red). Scale bars 200µm.

To further investigate the biological pathways involved in the induction of fibrosis in Cluster 2 patients, we assessed the expression of fibrosis-related genes in the patient’s lungs. We observed an increased expression of genes related to fibrinolysis, inflammatory cytokines, nitric oxide-induced, extracellular matrix deposition, and extracellular matrix remodeling in patients belonging to Cluster 2 (**Figure 5D**). A volcano plot indicates genes differentially expressed in samples from Cluster 2 compared to Cluster 1, including *Plg* (**Figure 5E**)*, Plat* (**Figure 5F**)*, Plau* (**Figure 5G**); *Ccr2* (**Figure 5H**)*, Ccl3* (**Figure 5I**); *Cdkna1* (**Figure 5J**)*; Serpina1* (**Figure 5K**)*, Timp2* (**Figure 5L**) and *Fgf1* (**Figure 5M**).

To further illustrate the differences between fibrosis and thrombosis in patients from these different clusters, we performed Masson-Goldner staining to assess erythrocytes and clot formation (bright red) and collagen deposition (green) in Patients’ lungs. By assessing three patients from each cluster, we found an increased collagen deposition in Cluster 2 patients, suggesting an increased fibrotic process (**Figure 5N**). By contrast, samples from Cluster 1 show increased clot formation, suggesting increased thrombotic processes (**Figure 5O**). Together, these data support the hypothesis that fatal cases of COVID-19 progress via induction of disseminated intravascular coagulation, culminating in a thrombotic process or via a fibrotic process, which is aggravated by the inflammasome-induced exacerbated inflammation.

## Discussion

COVID-19 is significantly lethal in nonvaccinated individuals, and although inflammation and cytokine storm are associated with poor clinical outcomes, the mechanisms underlying dysregulated inflammatory processes are unknown. The revelation that exacerbated inflammasome activation contributes to COVID-19 pathology (Junqueira et al., 2022; Rodrigues et al., 2021; Sefik et al., 2022) advanced the understanding of disease pathology, but a significant proportion of the lethal cases progresses with lower inflammasome activation. Our analysis of 47 fatal COVID-19 cases allowed for the classification of the patients into two groups. Cluster 2 showed a remarkably high inflammasome activation and hyperexpression of inflammatory genes with increased pulmonary fibrosis and worsened respiratory functions. By contrast, patients belonging to Cluster 1 died faster, with higher viral loads, reduced inflammatory process, and increased disseminated intracellular coagulation. Our data reveal two distinct profiles in lethal cases of COVID-19, thus indicating that the balance of viral replication and inflammasome-mediated pulmonary inflammation led to different clinical outcomes.

Patients belonging to Cluster 2, died of poor respiratory functions and increased fibrosis induced by the excessive inflammatory process. Inflammatory cytokines released upon inflammasome activation have been previously linked to the development of pulmonary fibrosis (Cho et al., 2020; Hussain et al., 2014; Lv et al., 2018; Meng et al., 2015; Meng et al., 2019; Sun et al., 2016). In addition, cytokines such as IL-1β, IL-18, and IL-1α have been described as triggering the activation of fibroblasts and stimulating the synthesis and accumulation of type I collagen, TIMP, collagenase, and PGE2 (Hussain et al., 2014; Postlethwaite et al., 1988), which may contribute to the triggering of the deleterious effects of inflammasomes in patients’ lungs. Interestingly, patients from Cluster 2 died with low viral loads and higher inflammasome activation; in many patients from this cluster, viral loads were not detected, suggesting that SARS-CoV-2 per see may not be required for continuous inflammasome activation in the lungs of these patients. How NLRP3 inflammasome is activated in the tissues is still unknown, it is possible that Damage-Associated Molecular Patterns (DAMPs) participate in this process. Alternatively, it is possible that viral proteins produced before viral elimination or undetectable levels of SARS-CoV-2 present in the tissues are sufficient to activate NLRP3 inflammasome. Future studies will be required to clarify these questions. We also tested whether the mechanical ventilation used in COVID-19 patients would interfere with inflammasome activation. This hypothesis is supported by data indicating that mechanical ventilation-induced hyperoxia can induce potassium efflux through the P_2_X_7_ receptor, thus leading to inflammasome activation and the secretion of proinflammatory cytokines (Dolinay et al., 2012; Jones et al., 2014; Kolliputi et al., 2010; Kuipers et al., 2012; Wu et al., 2013; Zhang et al., 2014). This does not appear to be the case in our study, as when we separated the COVID-19 patients into two groups according to the use or non-use of mechanical ventilation, we detected no differences in histopathological analyses and inflammasome activation (**Table 4**). Moreover, we confirmed that patients who underwent mechanical ventilation had a longer illness time, higher hypertension, less CRP and albumin, higher amounts of urea, and worsened pulmonary function (according to the PaO2/FiO2 and A-an O2 gradients). Thus, our data do not support the hypothesis that mechanical ventilation is directly associated with inflammasome activation.

**Table 4.** Characteristics of COVID-19 patients who were submitted or not to mechanical ventilation (MV).

| Demographics Characteristics | MV+<br>Mean (±SD) or<br>n (%) | MV-<br>Mean (±SD) or<br>n (%) | p value |
| --- | --- | --- | --- |
| N | 36 | 11 |  |
| Sex |  |  |  |
| Female | 17 (47.22%) | 6 (54.54%) | 0.7400* |
| Male | 19 (52.77%) | 5 (45.45%) |  |
| Age (years) | 65.61 (±13.52) | 75.72 (±17.61) | 0.0493** |
| Illness Time (days) | 20.94 (±10.76) | 8.72 (±5.40) | 0.0007** |
| ICU | 36 (100.00%) | 0 (0.00%) | >0.9999* |
| **Comorbidities** |  |  |  |
| Hypertension | 23 (63.88%) | 3 (27.27%) | 0.0433* |
| BMI | 30.61 (±8.52) | 34.56 (±14.46) | 0.4324** |
| Diabetes | 16 (44.44%) | 2 (18.18%) | 0.1644* |
| History of smoking | 11 (30.55%) | 3 (27.77%) | >0.9999* |
| Heart disease | 11 (30.55%) | 1 (9.09%) | 0.2440* |
| Lung disease | 9 (25.00%) | 2 (18.18%) | >0.9999* |
| Kidney disease | 6 (16.66%) | 1 (9.09%) | >0.9999* |
| History of stroke | 4 (11.11%) | 3 (27.77%) | 0.3296* |
| Autoimmune diseases | 1 (2.77%) | 1 (9.09%) | 0.4172* |
| **Laboratorial findings** |  |  |  |
| CRP (mg/dL) | 9.99 (±7.83) | 16.02 (±9.29) | 0.0377** |
| D-Dimers (µg/mL) | 4.99 (±4.68) | 4.56 (±3.62) | 0.7989** |
| LDH (mmol/L) | 5.78 (±5.76) | 2.49 (±1.13) | 0.0680** |
| Creatinine (mg/dL) | 1.58 (±1.39) | 1.05 (±0.50) | 0.2242** |
| Urea (mg/dL) | 138.18 (±54.07) | 64.48 (±34.12) | 0.0001** |
| AST (IU/L) | 109.61 (±98.16) | 84.81 (±54.14) | 0.4725** |
| ALT (IU/L) | 79.02 (±58.67) | 45.38 (±26.97) | 0.1254** |
| AST/ALT | 1.6 (±1.74) | 2.2 (±0.88) | 0.3542** |
| PT (INR) | 1.53 (±0.91) | 2.82 (±4.24) | 0.0900** |
| Albumin (g/dL) | 2.99 (±0.54) | 3.78 (±0.41) | 0.0034** |
| Blood Glucose (mg/dL) | 198.22 (±93.77) | 181.11 (±102.34) | 0.6340** |
| **Respiratory status** |  |  |  |
| Temperature (°C) | 37.51 (±1.79) | 36.99 (±1.53) | 0.3890** |
| P<sub>a</sub>O<sub>2</sub> (mmHg) | 79.57 (±20.19) | 53.99 (±24.12) | 0.0014** |
| Venous saturation (S<sub>v</sub>O<sub>2</sub>) | 70.86 (±14.36) | 50.71 (±31.26) | 0.0092** |
| P<sub>a</sub>O<sub>2</sub>/FiO<sub>2</sub><sup>#</sup> | 132.63 (±67.94) | 257.11 (±114.86) | <0.0001** |
| A-a O<sub>2</sub> Gradient Value<sup>#</sup> | 351.09 (±183.40) | 45.69 (±26.55) | <0.0001** |
| Respiratory Rate (mov/min) | 27.61 (±6.17) | 23.09 (±7.89) | 0.0669** |
| **Histopathological findings** |  |  |  |
| Fibrosis (% of area) | 30.28 (±14.05) | 29.09 (±12.21) | 0.8015** |
| Organizing Pneumonia (% of area) | 26.11 (±13.56) | 20.00 (±11.54) | 0.4520** |
| Acute Fibrinous and Organizing Pneumonia (% of area) | 19.50 (±12.48) | 26.00 (±15.16) | 0.3906** |
| Diffuse Alveolar Damage (% of area) | 28.82 (±19.34) | 43.75 (±16.85) | 0.0742** |
| Pneumonitis (% of area) | 23.52 (±17.33) | 20.00 (±8.16) | 0.5435** |
| Pulmonary infarction (% of area) | 25.00 (±10.00) | 10.00 (±0.00) | 0.2235** |
| **Inflammasome activation** |  |  |  |
| NLRP3 puncta per parenchyma area (mm<sup>2</sup>) | 204.3 (±155.5) | 204.0 (±156.6) | 0.9310** |
| ASC puncta per parenchyma area (mm<sup>2</sup>) | 251.8 (±192.5) | 267.3 (±133.1) | 0.4196** |

\* Fisher's exact test
\*\* t-Test

The mechanisms underlying dysregulated inflammatory processes and the cell types involved in inflammasome activation in COVID-19 are largely unknown. Macrophages are possibly the most effective cell types that trigger inflammasome activation (Broz and Dixit, 2016); according to this information, our data showed that macrophages in the lungs of COVID-19 patients are effectively infected and strongly induce inflammasome activation. These data are in agreement with previously published articles indicating high levels of viral RNA in lung monocytes and macrophages of COVID-19 patients (Delorey et al., 2021; Pontelli et al., 2022), as well as pronounced inflammasome activation in macrophages from COVID-19 patients (Junqueira et al., 2022; Rodrigues et al., 2021; Sefik et al., 2022). In addition to inflammasome activation in macrophages, the analysis of inflammasome activation in different pulmonary cell types indicates that endothelial cells exposed to SARS-CoV-2 express inflammasome proteins and contain active NLRP3/ASC puncta, thus demonstrating inflammasome activation. Despite reports indicating that endothelial cells are not productively infected by SARS-CoV-2 (Schimmel et al., 2021), precursors of hematopoietic and endothelial cells stimulated with SARS-CoV-2 spike protein increase the expression of *AIM2*, *NLRP1*, *NLRP3*, *IL1Β*, and *ASC* and trigger caspase-1 activation (Kucia et al., 2021; Ratajczak et al., 2021). In addition, inflammasome activation in endothelial cells has been previously reported (Paul et al., 2021; Xiang et al., 2011; Xu et al., 2013; Yang et al., 2016). These observations are consistent with our data indicating the expression of inflammasome components and inflammasome activation in CD34+ cells in the lungs of COVID-19 patients. Although these cells were stained positive for the SARS-CoV-2 spike protein, it is unknown whether infection and viral replication are required for inflammasome activation in these cells. It is possible that exosomes, released from COVID-19-infected cells, trigger NLRP3 inflammasome in endothelial cells (Sur et al., 2022).

Alternatively, It is possible that spike stimulation is sufficient for inflammasome activation, which is a feature that would explain reports indicating damage and dysfunction of these cells during COVID-19 and the characterization of COVID-19 as an endothelial disease (Hottz et al., 2020; Libby and Lüscher, 2020; Liu et al., 2021; Nuovo et al., 2021; Varga et al., 2020; Ward et al., 2021). Importantly, vascular endothelium is actively involved in the regulation of inflammation and thrombus formation. This is particularly important in COVID-19 because the interplay between inflammasome activation in macrophages and the endothelium can lead to pyroptotic macrophages releasing tissue factor (TF), which is an essential initiator of coagulation cascades that are frequently observed in severe cases of COVID-19 (Campos et al., 2021; Wu et al., 2019; Zhang et al., 2021). These data support the association of inflammasomes with the coagulopathy that is observed in COVID-19.

In this study, we demonstrated specific cell types involved in inflammasome activation in patients’ lungs and identified two distinct profiles in lethal cases of COVID-19. The revealed balance of viral-induced intravascular coagulation versus inflammasome-mediated pulmonary inflammation contributes to our understanding of disease pathophysiology and may contribute to decisions between immune-mediated or antiviral-mediated therapies for the treatment of critical cases of COVID-19.

## Methods

### Samples and Study Approval

Minimally invasive autopsy was performed on 47 patients diagnosed with SARS-CoV-2 at the Hospital das Clínicas da Faculdade de Medicina de Ribeirão Preto da Universidade de São Paulo, Brazil (Ribeirão Preto, SP, Brazil) from April to July, 2020, by Serviço de Patologia (SERPAT). Minimally invasive autopsy was done at bedside through post-mort surgical lung biopsy by a matching 14-gauge cutting needle (Magnum Needles, Bard) and a biopsy gun (Magnum, Bard). Moreover, a 3 cm incision on the more affected side of the chest between the fourth and fifth ribs were also used to provide extra lung tissue. All tissue samples were embedded in paraffin and fixed in formalin (Formalin-Fixed Paraffin-Embedded, FFPE). Lung tissue from biopsy of Lung Adenocarcinoma patients (n=5) were obtained from SERPAT. The project was approved by the Research Ethics Committee of FMRP/USP under protocol n° 4,089.567.

### Immunohistochemistry (IHC)

Tissue section in paraffin blocks were tested by immunohistochemistry using antibodies for the detection of NLRP3 (clone D2P5E; 1:3,000; Cell Signaling), ASC (1:2,000; Adipogen AL177), SFTPC (1:200, ThermoFisher PA5-71680), CD68 (1:200, Dako M0814), CD34 (1:500, Zeta Z2063ML), PDPN (1:200, ThermoFisher 14-9381-82), Anti-cleaved N-terminal GSDMD (1:200, Abcam ab215203), Caspase-1 (1:200, Abcam ab207802), IL-1β (1:200, Abcam ab2105) for the *in situ* detection of these inflammasome proteins. The Sequential Immunoperoxidase Labeling and Erasing (SIMPLE) technique was used to evaluate all markers in the same tissue, as previously published (Glass et al., 2009). Briefly, after incubation with primary antibody (overnight at 4°C), slides were incubated with immune peroxidase polymer anti-mouse visualization system (SPD-125, Spring Bioscience, 345 Biogen) and with chromogen-substrate AEC peroxidase system kit (SK-4200, 346 Vector Laboratories, Burlingame, CA). After high resolution scanning by a VS120 Olympus microscope, the coverslips were removed in PBS and the slides were dehydrated in an ethanol gradient to 95% ethanol. The slides were incubated in a series of ethanol to erase the AEC marking. Afterwards the slides were rehydrated and the antibodies were removed with an incubation for 2 min in a solution of 0.15 mM 351 KMnO4/0.01 M H_2_SO_4_, followed immediately by a wash in distilled water. Tissues were then remarked.

### Immunofluorescence

The slides were incubated with the primary antibodies, rabbit anti-human NLRP3 mAb (clone D2P5E; 1:300; Cell Signaling), rabbit anti-human ASC polyclonal antibody (1:200; Adipogen AL177), SFTPC (1:200, ThermoFisher PA5-71680), CD64 (1:200, Biolegend 139304), CD34 (1:500, Zeta Z2063ML), PDPN (1:200, ThermoFisher 14-9381-82), overnight at 4°C and with the secondary antibodies Goat anti-mouse Alexa fluor-647 (Invitrogen) or Goat anti-rabbit Alexa fluor-594 (Invitrogen). Images were acquired by the Axio Observer system combined with the LSM 780 confocal device microscope at 63x magnification (Carl Zeiss).

### Histological Evaluation

Paraffin-embedded lung tissues sections (3 μm) were stained by standard hematoxylin and eosin (H&E). Morphological lung injury patterns were evaluated by specialized pulmonary pathologists (ATF) blinded to clinical history. They were classified as absent or present with its extent of lung injury area by 5% cut-offs in Fibrosis, Organizing Pneumonia (OP), Acute Fibrinous and Organizing Pneumonia (AFOP), Diffuse Alveolar Damage (DAD), Cellular Pneumonitis, Thrombus Formation.

### RNA extraction and Real-Time Polymerase Chain Reaction for inflammatory genes

Total RNA from fresh lung tissue of SARS-CoV-2 patients and controls was obtained using Trizol reagent, and purification was performed according to the manufacturer’s instructions. The RNA was quantified by spectrophotometry in a NanoDrop 2000c spectrophotometer. The concentration was adjusted to 1 μg/µL, and the RNA was stored at −70 ◦C until reverse transcription. For inflammatory genes qPCR the total RNA was transcribed into complementary DNA (cDNA) using a High-Capacity cDNA Reverse Transcription kit (without an inhibitor) according to the protocol provided by the manufacturer (Thermo Fisher, Carlsbad, CA, USA). The reaction was prepared in a final volume of 20.0 µL containing 4.2 µL of H_2_O, 2.0 µL of buffer, 2.0 µL of random primers, 0.8 µL of dNTP Mix (100 mM), 1.0 µL of reverse transcriptase (RT) enzyme and 1 µL of RNA (1 ug/µL). The solution was then placed into a thermocycler with the following program: 25 ◦C for 10 min, 37 ◦C for 120 min and 85 ^◦^C for 5 min. The real-time PCR was performed in 96-well plates using Sybr Green reagents (Applied Biosystems, Waltham, MA, USA) and a Quant studio real-time PCR system (Applied Biosystems, Foster City, CA, USA). The real time-RT-PCR was carried out in a total volume of 20 μl on a 96-well MicroAmp Fast Optical plate (Applied Biosystems). Each well contained 10 μl SYBR Green qPCR Master Mix (Thermofisher), 1 μl of each primer (**Supplementary Table 1**), 2 μl cDNA (20 ng) and 7 μl RNase free water using the following protocol: initial denaturation at 95°C for 10 min, 40 cycles of denaturation at 95°C for 15 s followed by annealing/extension at 60°C for 60 s. Each PCR was followed by a dissociation curve analysis between 60-95°C. The Ct values were analyzed by the comparative Ct (ΔΔCt) method and normalized to the endogenous control GAPDH. Fold difference was calculated as 2^−ΔΔCt^

### Real-Time Polymerase Chain Reaction for Viral RNA

Detection and quantification of SARS-CoV-2 genes was performed with primer-probe sets for 2019-nCoV_N2 and gene E, according to US Centers for Disease Control and Prevention (Lu et al., 2020) and Charité group protocols (Corman et al., 2020). The genes evaluated (N2, E, and RNase-P housekeeping gene) were tested by one-step real-time RT-PCR using total nucleic acids extracted with TRIzol (Invitrogen). All real-time PCR assays were done on a Quant studio real-time PCR system (Applied Biosystems, Foster City, CA, USA). A total of 70 ng of RNA was used for genome amplification, adding specific primers (20 µM), and probe (5 µM), and with TaqPath 1-Step quantitative RT-PCR Master Mix (Applied Biosystems), with the following parameters: 25°C for 2 min, 50°C for 15 min, and 95°C for 2 min, followed by 45 cycles of 94°C for 5 s and 60°C for 30 s. Primers used were the following: N2 forward: 5′-TTACAAACATTGGCCGCAAA-3′, N2 reverse: 5′-GCGCGACATTCCGAAGAA-3′; N2 probe: 5′-FAM-ACAATTTGCCCCCAGCGCTTCAG-BHQ1-3′ (Lu et al., 2020); E forward: 5′-ACAGGTACGTTAATAGTTAATAGCGT-3′, E reverse: 5′-ATATTGCAGCAGTACGCACACA-3′; E probe: 5′-AM-ACACTAGCCATCCTTACTGCGCTTCG-BHQ-1-3′ (Corman et al., 2020); RNase-P forward: 5′-AGATTTGGACCTGCGAGCG-3′, RNase-P reverse: 5′-GAGCGGCTGTCTCCACAAGT-3′; and RNase-P probe: 5′-FAM-TTCTGACCTGAAGGCTCTGCGCG-BHQ-1-3′ (Lu et al., 2020). A plasmid of N2 protein and E protein was used for a standard curve construction for viral load quantification.

### Chest computed tomography images acquisition and evaluation

Chest x-radiography (CXR) and computed tomography (CT) exams were performed as part of the routine clinical evaluation. Chest radiographies were performed in conventional equipment, mainly in the anteroposterior incidence. CT images were performed in multidetector scanners (Brilliance CT Big Bore 16 - Philips, Holland, or Aquilion Prime 160 - Toshiba, Japan), using similar protocols for the acquisition of high-resolution images of the lungs (Raghu et al., 2018). Patients were scanned in the supine position without the administration of intravenous contrast media. Typical acquisition parameters were: 120 kVp tube voltage, 100–140 ref mAs (Koenigkam-Santos et al., 2022), 0.3–0.7 s gantry rotation time, reconstruction matrix size of 512×512, slice thickness, and increment of 1.0 mm, using standard (soft) and hard kernel filters. Imaging exams were independently evaluated by two thoracic radiologists (MKS and DTW), blinded to clinical data, laboratory, and pathology results as described (Koenigkam-Santos et al., 2022). Divergences were solved by consensus. All CXRs available were classified for the presence and grade of viral pneumonia (Revel et al., 2020). Pulmonary disease evolution on imaging was evaluated considering all exams from the initial to the last image before death. Tomographic images were evaluated similarly to CXR images.

### Statistical Analysis

For puncta quantification, all histological sections were viewed on a 63x objective for digitalizing random images using the LSM 780 system in the Axio Observer microscope, covering an area of about ∼1.7 mm^2^ of lung parenchyma analyzed per case. Manual counting of puncta and cells was blinded and performed using the acquired images. Morphometric analyzes were performed as described (Weibel, 1963). The quantification of expression by immunohistochemistry was performed by calculating the percentage of marked area, the scanned images were opened in the ImageJ software, using the IHQ Toolbox plugin, which consists of a semi-automatic color selection tool that selects the pixels positive for immunohistochemical marking, differentiating them from background and H&E marking, after selecting the positive pixels, the images were transformed into 8-bits and the area occupied by these tones was calculated. The distribution of the gene expression and biochemical marker and puncta count data was evaluated using the Shapiro–Wilk test; the data were analyzed using non-parametric Kruskal-Wallis, Mann-Whitney test and Spearman correlation. The data as violin plot graphs show median and quartiles. Statistical analyzes were performed using the GraphPad PRISM 5.0 program, with p<0.05 being considered statistically significant. Heatmaps were constructed using the heatmap.2 function in the R program (Project for Statistical Computing, version 3.4.1), the hierarchical clustering method used a correlation distance measure with ward.D2 and Canberra analysis.

## Data Availability

All data produced in the present study are available upon reasonable request to the authors

## Acknowledgments

We would like to thank Maira Nakamura, Amanda Zuin and Dr. Roberta Sales for technical support.

---

**Supplementary Table 1.**
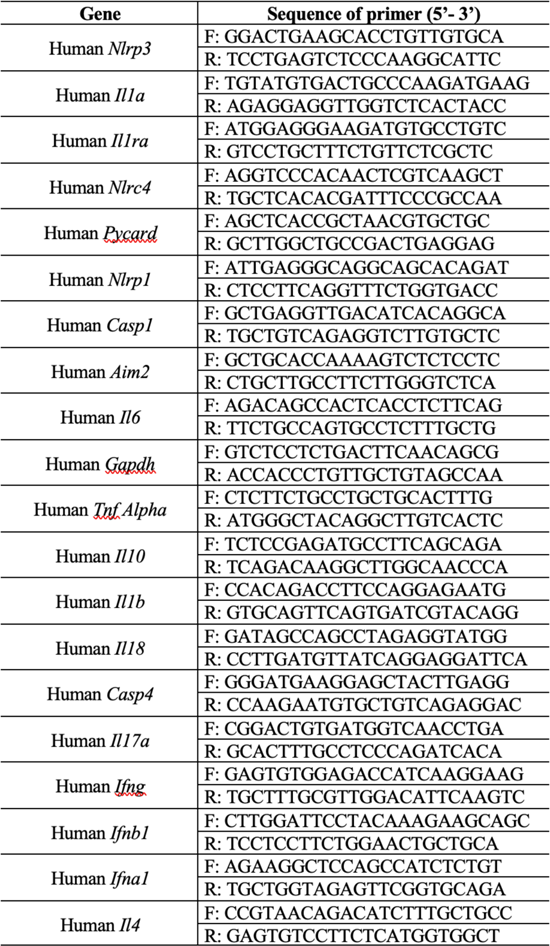
The list of primer sequences for real time-PCR

## Supplementary Figures and Supplementary Figure Legends

**Supplementary Fig. 1.**
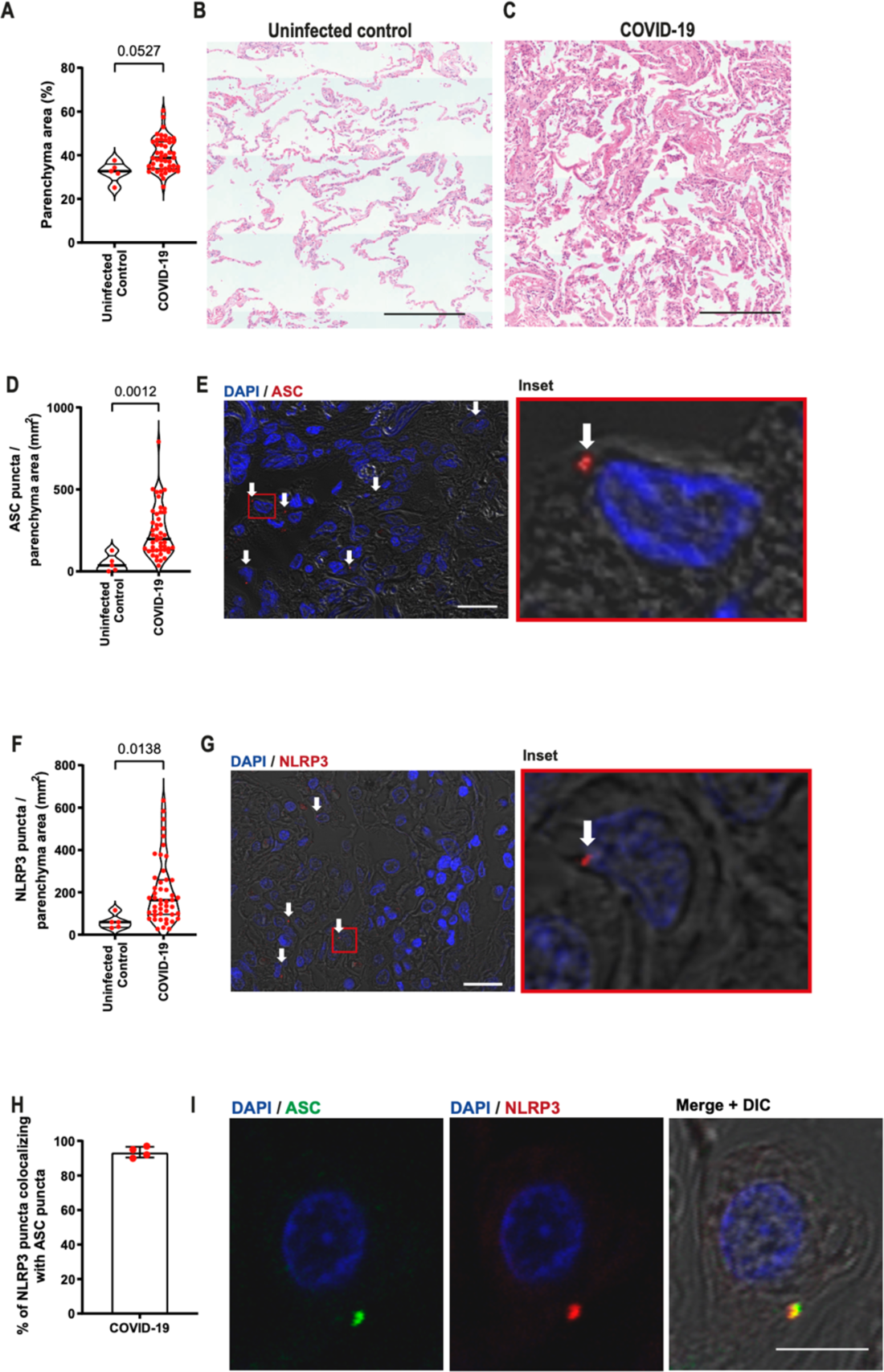
Histopathological patterns and inflammasome activation in lung autopsy of COVID-19 patients. Histopathological analysis of lethal cases of COVID-19 patients. (**A**) Proportion of lung parenchyma area (loss of airspace) of COVID-19 patients and uninfected controls (benign area of the lungs from adenocarcinoma patients). (**B-C**) Representative images of H&E stain showing the lung parenchyma. Scale bars 200 µm. (**D-G**) Multiphoton microscopy analysis of lung autopsies of 47 COVID-19 patients and 5 uninfected controls. Tissues were stained with anti-ASC (**D, E**) or anti-NLRP3 (**F, G**) for quantification of cells with inflammasome puncta in lung autopsies (in red, indicated by white arrows). DAPI stains cell nuclei (blue). Insets indicate a higher magnification of the indicated region (red rectangle). Scale bars 20 µm. (**H**) Percentage of NLRP3 puncta colocalizing with ASC puncta in the lungs of five COVID-19 patients. (**I**) Representative images showing ASC (green) and NLRP3 (red) colocalization. Scale bar 10 µm. Each dot in the figure represents the value obtained from each individual. P-values are shown in the figures comparing the indicated groups, as determined by Mann–Whitney test. Data are represented as violin plots with median and quartiles. The images were acquired by multiphoton microscope using a 63x oil immersion objective and analyzed using ImageJ Software.

**Supplementary Fig. 2.**
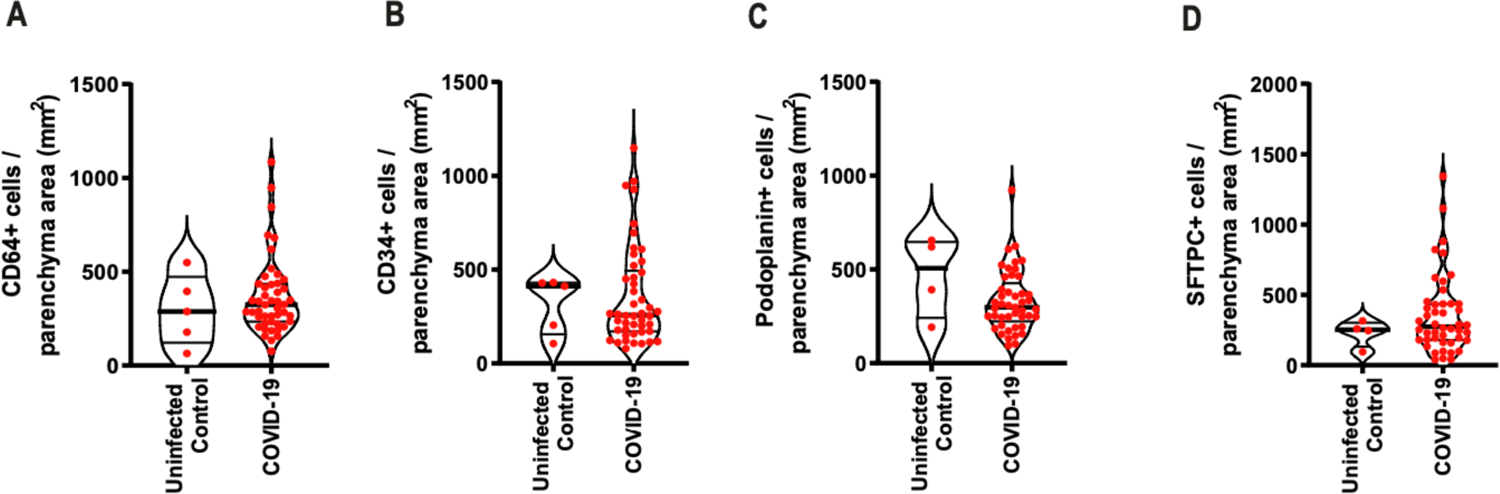
Total numbers of macrophages, endothelial cells, and type I and II pneumocytes are similar in the lungs of COVID-19 patients and uninfected controls. Multiphoton microscopy analysis of lung autopsies of 47 COVID-19 patients and 5 uninfected controls (benign area of the lungs from adenocarcinoma patients). (A-D) Numbers of macrophages (CD64^+^, A), endothelial cells (CD34^+^, B), type I pneumocytes (PDPN^+^, C) and type II pneumocytes (SFTPC^+^, D) per lung parenchyma area. Each dot in the figures represents the value obtained from each individual. P-values are shown in the figures comparing the indicated groups, as determined by Mann–Whitney test. Data are represented as violin plots with median and quartiles.

**Supplementary Fig. 3.**
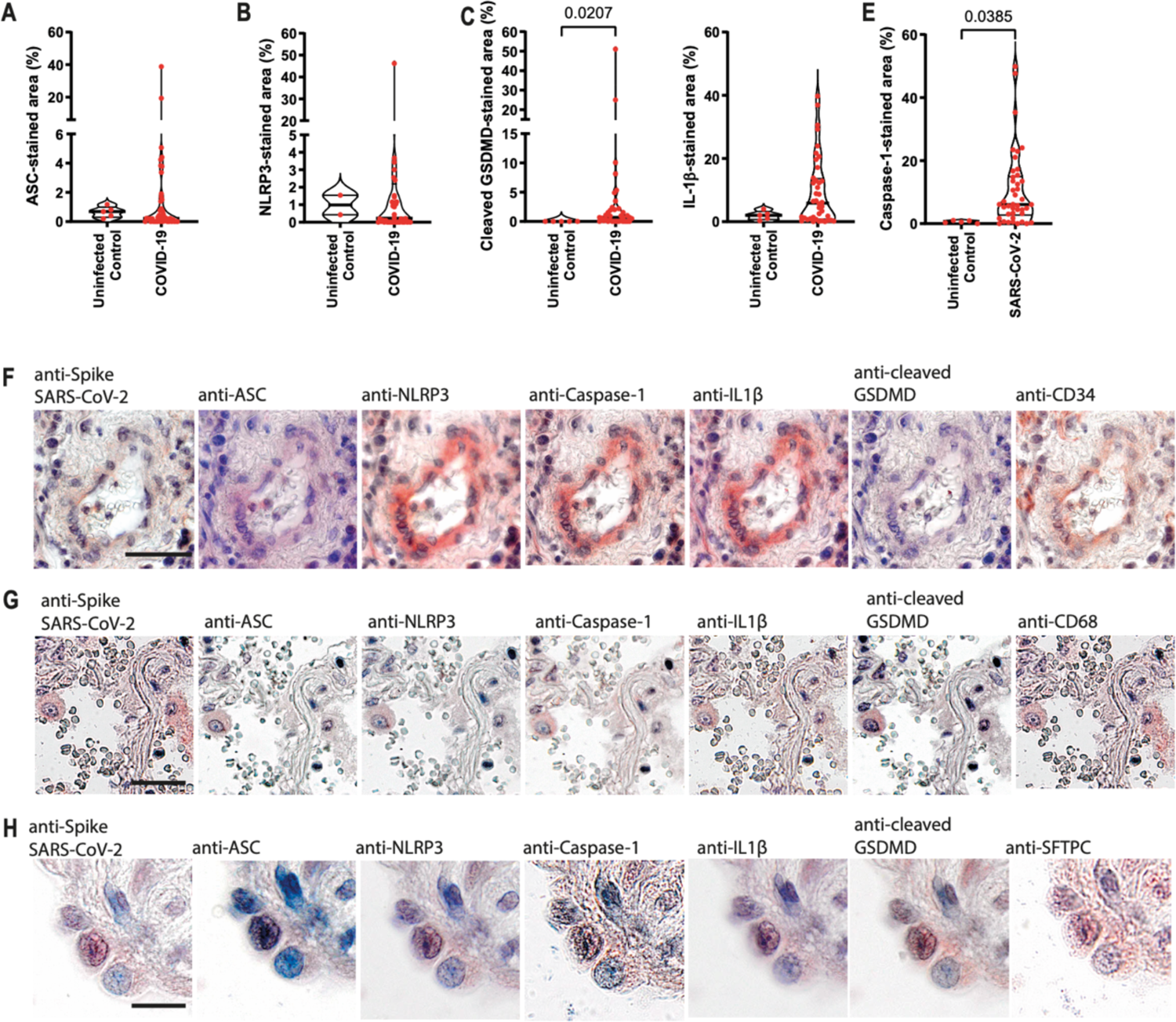
Immunohistochemistry analysis of lung autopsies of patients with COVID-19 and uninfected controls. Immunohistochemistry analysis of lung autopsies of 47 COVID-19 patients and 5 uninfected controls (benign area of the lungs from adenocarcinoma patients). Quantification of the percentage of area stained for ASC (**A**), NLRP3 (**B**), cleaved GSDMD (**C**), IL-1β (**D**) and Caspase-1 (**E**). Each dot in the figures represents the value obtained from each individual. P-values are shown in the figures comparing the indicated groups, as determined by Mann–Whitney test. Data are represented as violin plots with median and quartiles. (**F-H**) Representative images of lungs from fatal cases of COVID-19, indicating colocalization of viral Spike, ASC, NLRP3, Caspase-1, IL-1β and cleaved GSDMD in endothelial cells (CD34^+^, **F**), macrophages (CD68^+^, **G**), and type II pneumocytes (SFTPC^+^, **H**).

**Supplementary Fig. 4.**
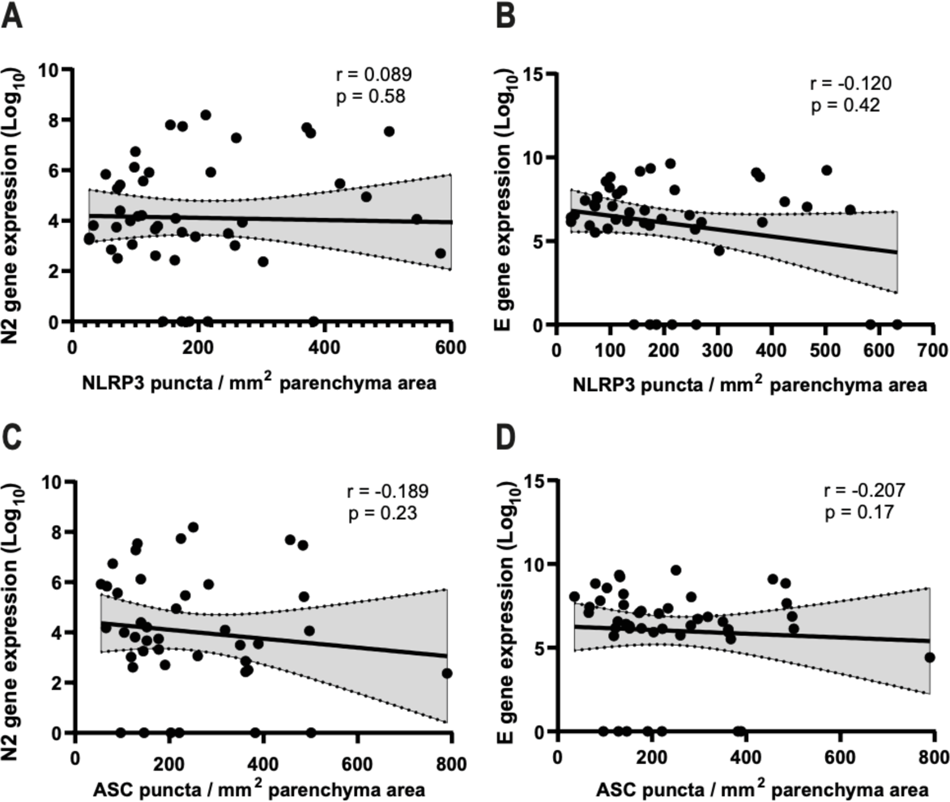
Non-significant correlation or viral loads and inflammasome activation in lethal cases of COVID-19 patients. Spearman correlation of pulmonary viral load and inflammasome activation in 47 fatal COVID-19 patients. (**A**) Correlation of viral N2 with NLRP3 puncta per parenchyma area; (**B**) Correlation of viral E with NLRP3 puncta per parenchyma area; (**C**) Correlation of viral N2 with ASC puncta per parenchyma area; (**D**) Correlation of viral E with ASC puncta per parenchyma area. r and p-value are indicated in the figure.

**Supplementary Fig. 5.**
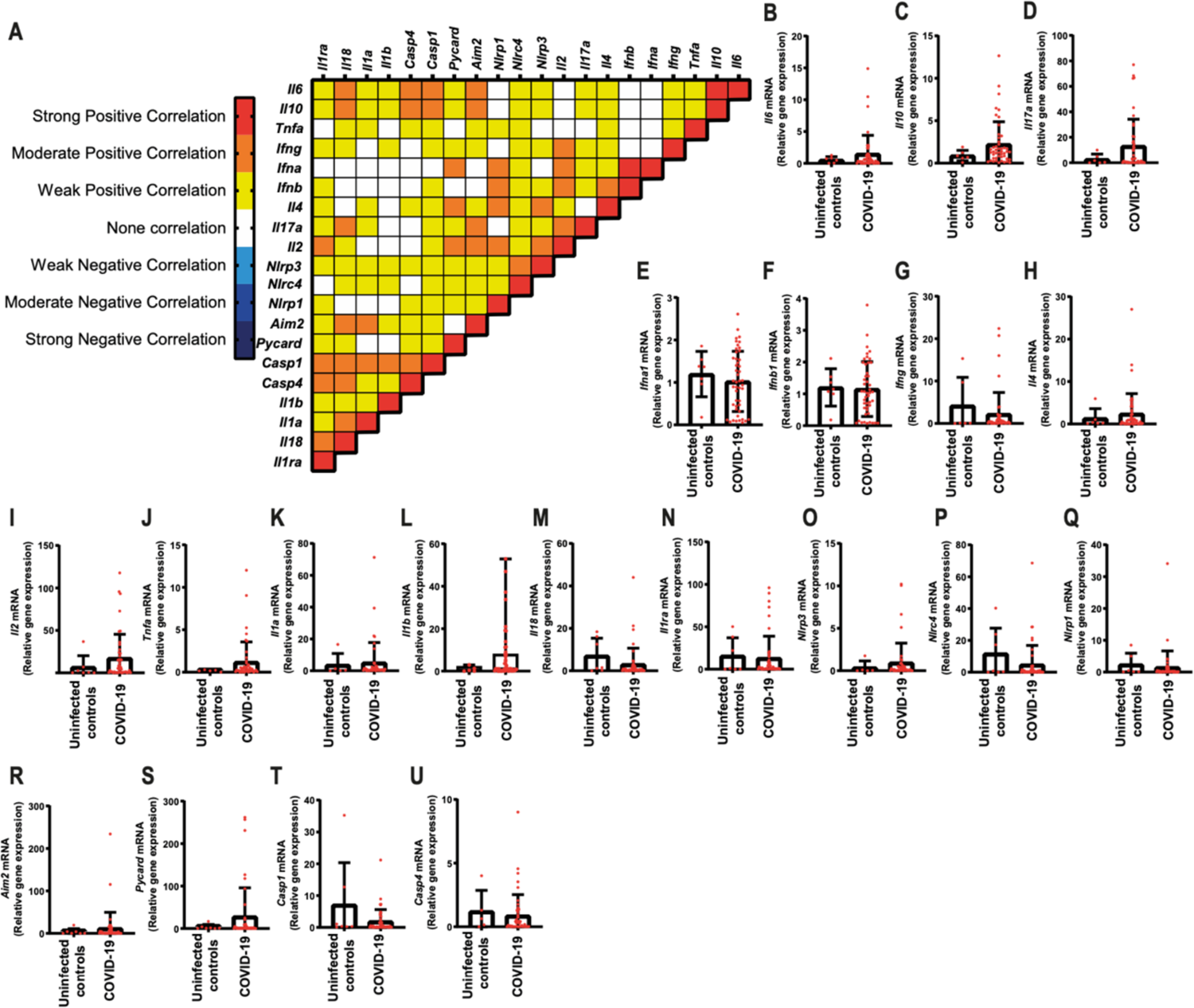
Gene expression in lungs of COVID-19 patients. Correlation matrix of inflammasome and inflammatory gene expression in lung autopsy of 47 COVID-19 patients (**A**). Colors indicate correlation scores, categorized as positive strong correlation (r ≥ 0.70; red); moderate positive correlation (0.50 ≥ r ≤ 0.70; orange); weak positive correlation (0.30 ≥ r ≤ 0.50; yellow); negative strong correlation (r ≥ −0.70; dark blue); negative moderate correlation (−0.50 ≥ r ≤ −0.70; blue) or negative weak correlation (−0.30 ≥ r ≤ −0.50; light blue). Only correlations with p<0.05 are represented in the correlation matrix. (**B-U**) Expression of mRNA in the lung autopsies of COVID-19 patients and uninfected controls (benign area of the lungs from adenocarcinoma patients). Selected genes were *Il6* (**B**), *Il10* (**C**), *Il17* (**D**), *Ifna1* (**E**), *Ifnb1* (**F**), *Ifng* (**G**), *Il4* (**H**), *Il2* (**I**), *Tnfa* (**J**), *Il1a* (**K**), *Il1b* (**L**), *Il18* (**M**), *I1ra* (**N**), *Nlrp3* (**O**), *Nlrc4* (**P**), *Nlrp1* (**Q**), *Aim2* (**R**), *Pycard* (**S**), *Casp1* (**T**), *Casp4* (**U**).

**Supplementary Fig. 6.**
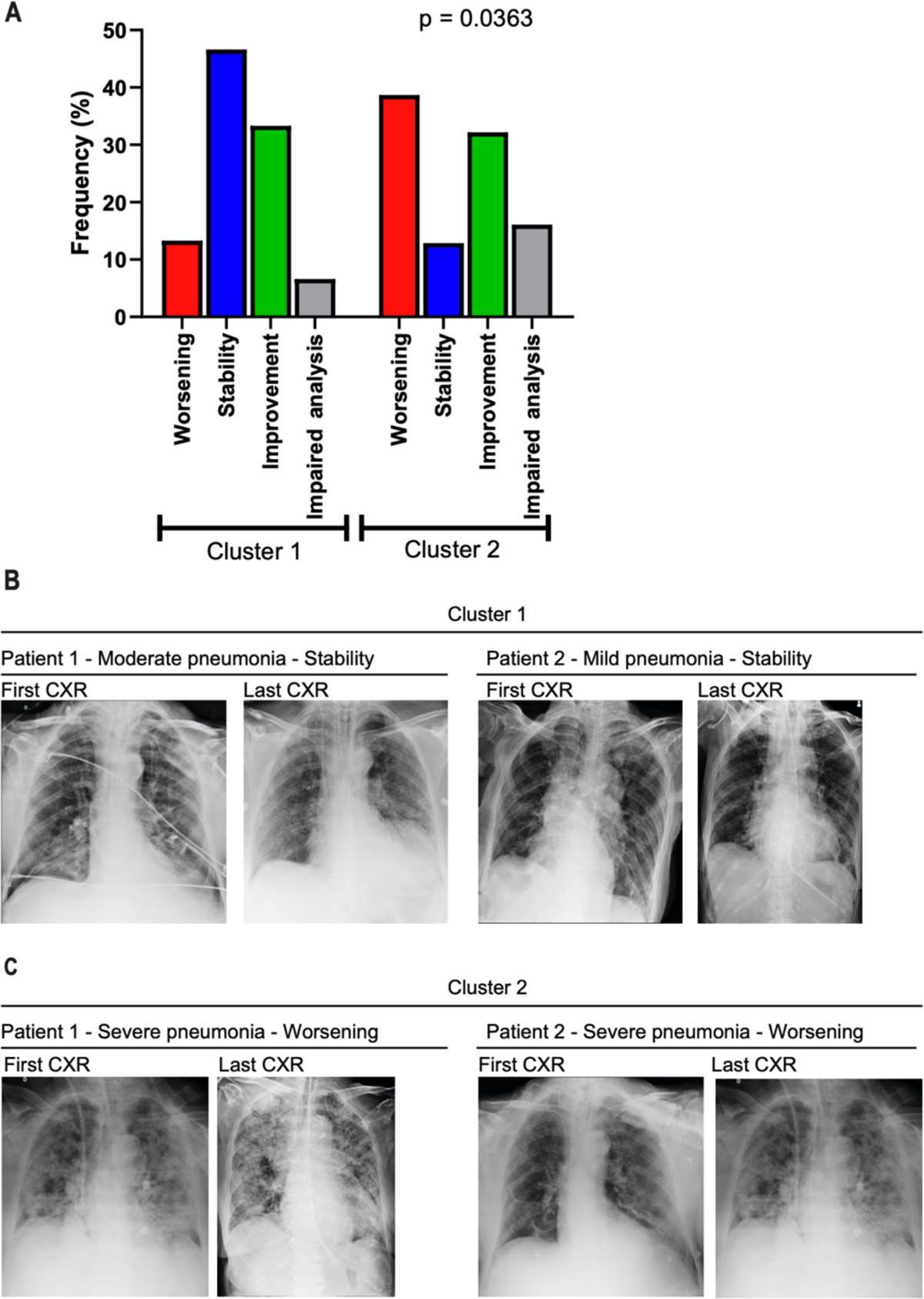
CXR evolution of patients from Cluster 1 and Cluster 2. Analysis of the first and last Chest x-radiography (CXR) of 47 COVID-19 patients belonging from Cluster 1 (n=15) and Cluster 2 (n=31). (**A**) patients with reduced opacities (green), stability (blue) and increased opacities (red) comparing the fists and first CXR. Impaired analyses are shown in gray. Representative images of first and last CXR from two patients from Cluster 1, indicating stability in moderate and mild pneumonia (**B**) and two patients from Cluster 2, indicating worsening conditions in cases of severe pneumonia (**C**).

